# Evaluating the Impact of Population-Based and Cohort-Based Models in Cost-Effectiveness Analysis: A Case Study of Pneumococcal Conjugate Vaccines in Infants in Germany

**DOI:** 10.1101/2025.02.21.25322688

**Authors:** Johnna Perdrizet, Dominik Schröder, Felicitas Kühne, Julia Schiffner-Rohe, Maren Laurenz, Christian Theilacker, Aleksandar Ilic, An Ta, Christof von Eiff

## Abstract

**Introduction:** Cost-effectiveness analysis (CEA) is crucial when evaluating the health and economic value of vaccines compared to the current standard of care (SoC) and provides essential information to assist decision-makers in maximizing health gains when allocating resources. The design of the CEA should address the specific policy questions, disease area, vaccine characteristics, and consider all relevant vaccination effects on the population.

**Areas covered:** We presented a case study on the CEA of pneumococcal conjugate vaccines (PCVs) in infants in Germany using a closed single cohort-based approach versus a population-based approach. Except for the design of the modelled population/cohort, all other inputs and characteristics were kept identical in the cost-effectiveness model. We contrasted model results, inferences, and conclusions between both design approaches.

**Expert Opinion:** CEA must carefully consider the included population in the analysis based on their specific policy questions and the characteristics of the vaccine being evaluated. The choice between population-based and closed single-cohort models fundamentally depends on whether the vaccine affects disease transmission dynamics. Population models are essential for vaccines that disrupt transmission patterns across population groups, such as PCVs in infants, while closed single-cohort models are suitable for vaccines impacting only vaccinated individuals without affecting disease transmission.

**Article highlights:**

- Identifying the appropriate model design is crucial for conducting cost-effectiveness analyses (CEAs) of vaccines, particularly when addressing vaccine technical committee (VTC) policy questions, which aim to optimize individual and population health benefits.
- Closed single cohort-based designs track a group of individuals, while population-based designs evaluate an entire cross-sectional population, making the choice between the two designs vital when vaccines have secondary, indirect effects.
- We presented a case study comparing PCV20 with PCV13 and PCV15 in infants in Germany using a closed single cohort-based approach and a population-based approach.
- Modelled results highlighted that the closed single cohort-based approach substantially underestimated public health benefits and economic advantages associated with PCV20, whereas the population-based approach demonstrated PCV20 as cost-saving strategy while offering superior health outcomes, indicating it as a dominant vaccination option when accounting for Germany’s entire population.
- Selecting an inappropriate model design for CEAs of vaccines could result in unintended consequences, such as adversely affecting national recommendations, policies, and programs, leading to suboptimal decision-making for population health.
- Researchers and policymakers must carefully select appropriate population frameworks and adhere to methodological guidelines to ensure accurate inferences in vaccine economic evaluations.

## 1 Introduction

Cost-effectiveness analysis (CEA) is crucial when evaluating the health and economic value of vaccines compared to the current standard of care (SoC) and provides essential information to assist decision-makers in maximizing health gains when allocating resources [1,2]. The modeling approach for a CEA should be tailored to the specific policy questions, disease epidemiology, and vaccine characteristics. Health gains for most vaccines are largely driven by indirect effects, where unvaccinated individuals are protected from the reduction of transmission within the population. Quantifying secondary benefits such as indirect effects can present methodological challenges in cost-effectiveness models [3].

The Standing Committee on Vaccination (STIKO) is Germany’s vaccine technical committee (VTC) that develops evidence-based vaccination recommendations for the population [4]. The overarching goal forming the policy questions of the STIKO is to maximize both individual- and population-level benefits, by balancing vaccine safety, efficacy and effectiveness while considering the cost-effectiveness of different vaccination strategies [5]. STIKO employs CEAs comparing new vaccines or vaccine schedules with the current standard of care (SoC), therapeutic interventions or scheduling strategies as part of their assessment [4,6,7]. Model outcomes evaluated include the impact of the novel intervention on the disease burden and associated costs for the entire country population [4,5].

To achieve this objective, CEA should consider all relevant vaccination outcomes, including direct effect for the vaccinated and, if any, indirect effects on the entire population. Accurate modeling of the targeted population is fundamental, especially for vaccines that affect diverse populations with varying characteristics and dynamics. The choice of model design for the population approach can considerably impact the model’s outcomes [8]. Different approaches offer unique capabilities in capturing population heterogeneity, demographic and epidemiological dynamics, and accounting for long-term intervention effects. While cohort design tracks individuals over time, population design evaluates the entire population within a specific geographical area at a given time.

Recent evaluations of the 20-valent pneumococcal conjugate vaccine (PCV20) have applied both model designs, highlighting their distinct applications in vaccine economic assessments. This vaccine is indicated for the prevention of invasive disease, pneumonia, and acute otitis media caused by *Streptococcus pneumoniae* in children and can provide an instructive case on how the population approach in CEA impacts vaccine assessment outcomes [9]. Two published CEA studies comparing PCV20 to a recommended SoC, the 13-valent PCV (PCV13) or the 15-valent PCV (PCV15) in Germany’s pediatric vaccination program used different population frameworks, leading to substantially different conclusions [10,11]. Therefore, the aim of this analysis is to compare the impact of a closed single-cohort population approach with a population single-cohort approach in a cost-effectiveness model, using PCV20 in Germany’s infant population as a case study.

## 2 Methods

A closed single-cohort and a population single-cohort Markov model were developed to estimate the cost-effectiveness for routine PCV. For a direct comparison between the two approaches, the cost-effectiveness model from Ta et al. which included assumptions and inputs on modelling perspective, cycle length, discount rate, vaccine effectiveness, resource use, costs, and benefits, was used [11]. The only modification made from the original publication was the adaptation of the population design approach.

### Included population

For the closed single-cohort model we used only one birth cohort consisting of 791,254 newborns. Conversely, the population single-cohort model used Germany’s population data from German Federal Statistical Office from the year 2022 (N= 83,772,723) stratified by age [12].

### Comparators

Both models compared PCV20 under a 3+1 vaccination schedule with PCV13 and PCV15, both administered using a 2+1 vaccination schedule.

### Outcomes

The CEA reported a full list of outcomes as reported in the original study by Ta et al including health outcomes (pneumococcal disease cases, stratified by invasive pneumococcal disease [IPD], hospitalized and non-hospitalized pneumonia and otitis media, number of deaths, life years [LYs] and quality-adjusted life years [QALYs]), economic or cost outcomes [in €] including cost of vaccination [doses and administration], medical costs, costs of sequelae, and societal cost of disease), and incremental results (incremental cost per LY and incremental cost per QALY).

## 3 Results

In the closed single-cohort model (N=791,254), which followed only one birth cohort over 10 years, PCV20 was associated with higher total costs compared to both PCV13 and PCV15 (Tables 1 & 2). Compared with PCV13, PCV20 resulted in incremental cost-effectiveness ratios (ICERs) of €15,948 per LY gained and €6,280 per QALY gained in the closed-single cohort model. PCV15 was associated with higher incremental costs than PCV20, with ICERs of €42,135 and €16,473 per LY and QALY gained, respectively.

**Table 1.**
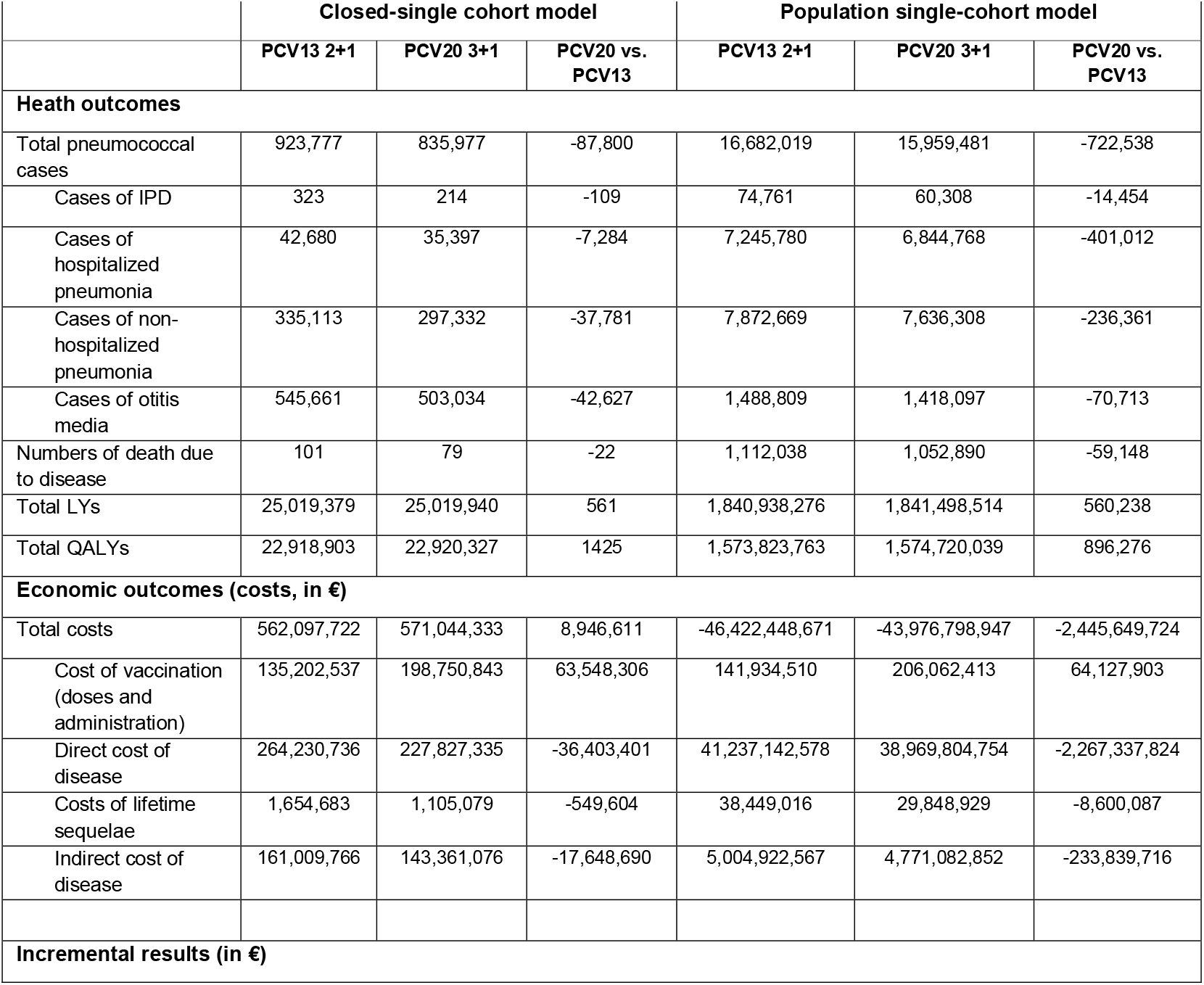
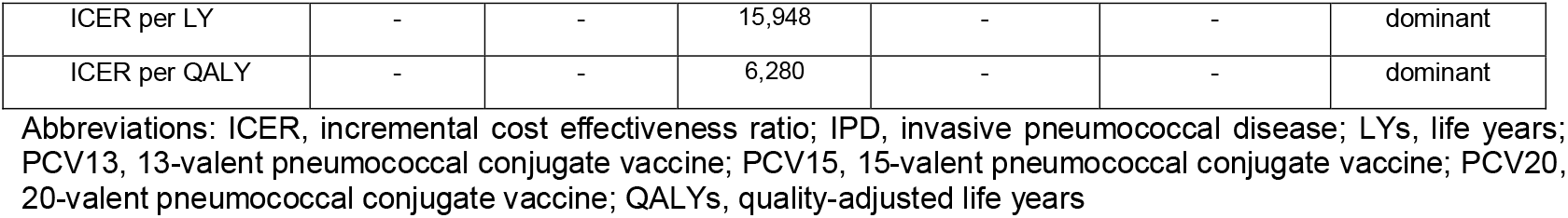
Cost-effectiveness results of PCV20 vs. PCV13 in closed single-cohort and population single-cohort models.

**Table 2.**
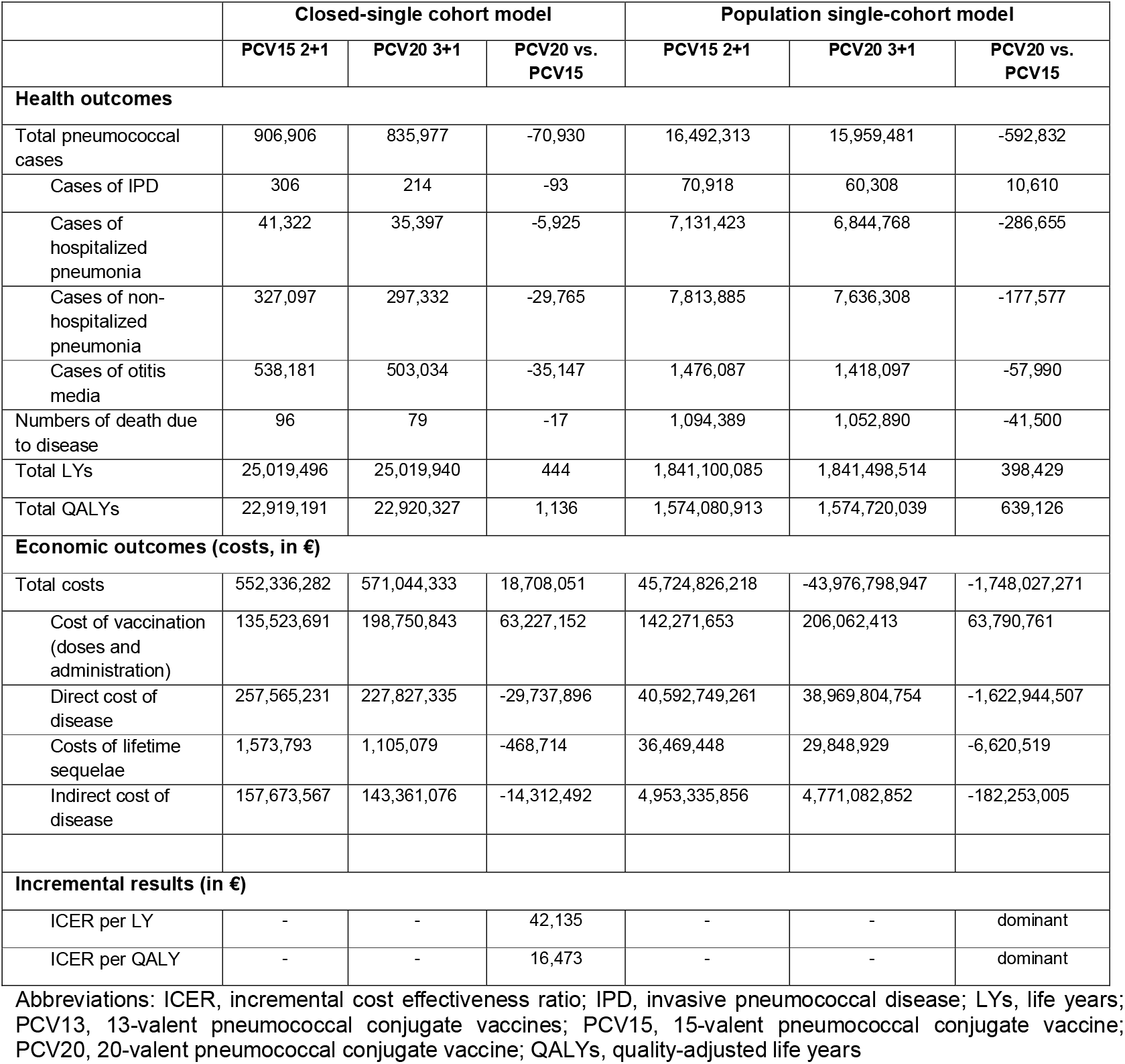
Cost-effectiveness results of PCV20 vs. PCV15 in closed single-cohort and population single-cohort models.

|  | Closed-single cohort model |  |  | Population single-cohort model |  |  |
| --- | --- | --- | --- | --- | --- | --- |
|  | PCV15 2+1 | PCV20 3+1 | PCV20 vs. PCV15 | PCV15 2+1 | PCV20 3+1 | PCV20 vs. PCV15 |
| **Health outcomes** |  |  |  |  |  |  |
| Total pneumococcal cases | 906,906 | 835,977 | -70,930 | 16,492,313 | 15,959,481 | -592,832 |
| Cases of IPD | 306 | 214 | -93 | 70,918 | 60,308 | 10,610 |
| Cases of hospitalized pneumonia | 41,322 | 35,397 | -5,925 | 7,131,423 | 6,844,768 | -286,655 |
| Cases of non-hospitalized pneumonia | 327,097 | 297,332 | -29,765 | 7,813,885 | 7,636,308 | -177,577 |
| Cases of otitis media | 538,181 | 503,034 | -35,147 | 1,476,087 | 1,418,097 | -57,990 |
| Numbers of death due to disease | 96 | 79 | -17 | 1,094,389 | 1,052,890 | -41,500 |
| Total LYs | 25,019,496 | 25,019,940 | 444 | 1,841,100,085 | 1,841,498,514 | 398,429 |
| Total QALYs | 22,919,191 | 22,920,327 | 1,136 | 1,574,080,913 | 1,574,720,039 | 639,126 |
| **Economic outcomes (costs, in €)** |  |  |  |  |  |  |
| Total costs | 552,336,282 | 571,044,333 | 18,708,051 | 45,724,826,218 | -43,976,798,947 | -1,748,027,271 |
| Cost of vaccination (doses and administration) | 135,523,691 | 198,750,843 | 63,227,152 | 142,271,653 | 206,062,413 | 63,790,761 |
| Direct cost of disease | 257,565,231 | 227,827,335 | -29,737,896 | 40,592,749,261 | 38,969,804,754 | -1,622,944,507 |
| Costs of lifetime sequelae | 1,573,793 | 1,105,079 | -468,714 | 36,469,448 | 29,848,929 | -6,620,519 |
| Indirect cost of disease | 157,673,567 | 143,361,076 | -14,312,492 | 4,953,335,856 | 4,771,082,852 | -182,253,005 |
| **Incremental results (in €)** |  |  |  |  |  |  |
| ICER per LY | - | - | 42,135 | - | - | dominant |
| ICER per QALY | - | - | 16,473 | - | - | dominant |

Abbreviations: ICER, incremental cost effectiveness ratio; IPD, invasive pneumococcal disease; LYs, life years; PCV13, 13-valent pneumococcal conjugate vaccines; PCV15, 15-valent pneumococcal conjugate vaccine; PCV20, 20-valent pneumococcal conjugate vaccine; QALYs, quality-adjusted life years

In contrast, the population single-cohort model, which included Germany’s entire population (N=83,772,723) demonstrated PCV20 as cost saving with better health outcomes compared to both PCV13 and PCV15. The results showed negative ICERs for both LY and QALY, indicating that PCV20 was the dominant strategy over both lower-valent alternatives.

## 4 Discussion

The aims of this case study were twofold: (1) to directly compare the impact of different population approaches on CEA results, and (2) to demonstrate the importance of selecting the appropriate model to address the VTC policy question. Despite using identical inputs, the conclusions drawn from the two modeling approaches differed. The population-based single-cohort model demonstrated PCV20 as the dominant vaccination strategy, while the closed single-cohort model estimated improved health outcomes associated with PCV20 at higher costs.

Indirect effects stemming from infant pediatric PCV programs (i.e. protection conferred to unvaccinated individuals of all ages) have been well-documented t [14-16]. As such, the closed single-cohort model for PCV in infants provided only limited insights into the policy questions posed by STIKO’s and other VTCs. This approach focused solely on a single birth cohort and failed to account for the broader population, thereby underestimating both health outcomes and economic benefits at the population level [4,13]. While this approach accounted for indirect effects within the single birth cohort it excluded indirect effects for the remaining approximately 99% of the existing population in Germany.

By contrast, the population single-cohort model incorporated both direct and indirect vaccine effects across the entire population. This approach provided more sufficient evidence to inform policy decisions, better aligned with the policy questions posed by STIKO and other VTCs regarding the implementation of PCV20 in infant vaccination program. Our analysis demonstrated that when accounting for underlying indirect effects, the population-based approach estimated greater disease prevention (i.e. more disease cases averted) at similar vaccination costs compared to the single-cohort approach. This led to a change in the qualitative conclusion, with PCV20 being dominant in the population-based approach, whereas it was considered cost-effective in the closed single-cohort model.

A related study by Standaert et al. compared a cohort- and population-based model designs to assess health effects and healthcare utilization of PCVs [17]. While Standaert et al. focused on different outcomes, the findings aligned with our analysis, concluding that a population-based approach should be used when an intervention, like PCVs in infants, is known to offer indirect effects across the entire population. This study, along with ours, reinforced the importance of selecting the appropriate model design to address the policy question effectively.

## 5 Conclusion

This study underscored the critical importance of accurately identifying the relevant population when conducting CEAs of vaccines for effectively addressing VTC policy questions. This is especially important when a vaccine produces indirect effects in individuals who are not directly vaccinated, given VTCs aim to optimize both individual and population-level health benefits. This case study comparing PCV20 to PCV13 and PCV15 in infants in Germany revealed that a closed single cohort-based approach significantly underestimated the potential public health benefits and economic advantages of introducing PCV20. Researchers performing economic evaluations of vaccines need to be especially careful when selecting the suitable population framework. Likewise, policymakers should be mindful of the potential misinterpretations arising from vaccine CEAs that do not adhere to proper methodological guidelines.

## 6 Expert Opinion

Health economic modelers must carefully consider modelling framework based on their specific policy questions and the characteristics of the vaccine being evaluated. The choice between a population-based and a closed-single-cohort model fundamentally depend on whether the vaccine disrupts disease transmission dynamics. The choice between these two population approaches is rarely discussed in economic evaluation studies of vaccines. Closed single cohort models are appropriate for vaccines that do not affect disease transmission and are only protective for vaccinated individuals. Tick-borne encephalitis vaccines can be used as an example where infection occurs mostly through tick bites and does not spread from human-to-human contact [19,20]. Another example is the recombinant zoster vaccine (RZV), which prevents shingles resulting from the reactivation of the dormant varicella-zoster virus within the individual and thus, does not influence disease transmission [21]. In contrast, population-based models are often essential for vaccines that disrupt transmission patterns across population groups, such as PCVs in infants. Another example is the original pertussis vaccine, which after it’s widespread availability in the 1940s demonstrated indirect effects by sharp reductions in both vaccinated and non-vaccinated infants and older populations [22]. Nonetheless economic analyses assessing the pertussis vaccine, like PCVs infants, do not always use the population approach including indirect effects [23]. For vaccines that produce indirect effects, the population-based approach is often necessary to answer the corresponding policy question, whilst a closed single-cohort approach might be suitable if researchers are only interested in a subset of the population (i.e., private market, immunocompromised or chronic medical conditions children) whereby secondary effects in the current existing population are not in focus of the policy question.

Our comparative analysis of PCV20 in Germany demonstrated how using an unsuitable model structure for addressing the policy question of VTCs could lead to different conclusions about cost-effectiveness of the intervention. Modelling a single birth cohort resulted in health benefits at higher incremental costs, while including the existing population led to both cost-savings and greater health benefits associated with PCV20 compared to PCV13 and PCV15. This difference in conclusions between the two model approaches was mainly attributed to the cohort model’s limitation in capturing indirect effects. Including indirect effects in closed-single-cohort models would underestimate the overall impact of vaccination programs, as these models typically focus on a single cohort in one dimension, tracking its members as they age and eventually die. Indirect effects, however, arise from interactions both within the vaccinated cohort and the remaining population, which can be accounted for in population-based models.

To advance vaccine economic modeling, it is essential to raise awareness about the critical importance of model choices in vaccine evaluations. Those who develop models need to be transparent on how different model structures capture or miss various effects of the vaccine intervention, rationale for the selected modelling approach, and how this choice could impact the results and inferences relevant for VTCs policy question. Our analysis indicated that relying on simple model approaches may overlook crucial benefits, generating imprecise results leading to faulty conclusions, and suboptimal decision-making. Additionally, CEA should strive to integrate the best available evidence while acknowledging data gaps and aligning model design with the local decision-making context. When reliable country-specific inputs are limited, leveraging data from other settings may be necessary, but the generalizability should be thoughtfully examined.

Looking ahead, vaccine economic evaluations will continue to provide valuable insights, even when some data remain unavailable. Newer vaccines, such as PCV20, respiratory syncytial virus vaccines, and COVID-19 vaccines, will benefit from the increasing availability of post-introduction real-world data, which is expected to enhance modelling by reflecting the evolving epidemiology of various diseases. These data will help to refine the estimates and allow for more precise assessments of the previously modeled impact. While cost-effectiveness is only one of many important considerations alongside vaccine safety, efficacy, and effectiveness, the growing competition for resources and the need to quantify the broader economic consequences of different vaccination strategies will likely increase the demand for locally relevant, scientifically rigorous, and policy-focused CEA. Ensuring that models adhere to decision-makers’ guidelines and justifications of any deviations throughout the model development process is critical for maintaining transparency and relevance, while appropriately addressing key areas of uncertainty.

## Statements/Declarations

### Funding details

Sponsorship for this study and accelerated publication fee were funded by Pfizer Inc.

### Declaration of interest

Johnna Perdrizet, Dominik Schröder, Felicitas Kühne, Julia Schiffner-Rohe, Maren Laurenz, Christian Theilacker, Aleksandar Ilic, Christof von Eiff are employees of Pfizer. An Ta is an employee of Cytel Inc., which received consulting fees from Pfizer Inc. for developing the cost-effectiveness model that was reanalyzed in this manuscript. An Ta contributed to this manuscript outside of her employment at Cytel, Inc. and received no funding.

### Data Availability Statement

All data generated or analyzed during this study are included in this published article.

### Author contributions

All the authors (J Perdrizet, D Schröder, F Kühne, J Schiffner-Rohe, M Laurenz, C Theilacker, A Ilic, A Ta, C von Eiff) (1) made substantial contributions to the study concept or the data analysis or interpretation, (2) drafted the manuscript or revised it critically for important intellectual content, (3) approved the final version of the manuscript to be published, and (4) agreed to be accountable for all aspects of the work.

### Ethical Approval

This article was based on previously conducted studies and does not contain any new studies with human participants or animals performed by any of the authors; as such, ethical approval was not required.

